# An Edge-AI Pose-Estimation Platform for Real-Time Posture and Posture-Transition Monitoring: A Feasibility Study for Pre-Emptive Fall-Risk Alerting in Elderly and Dementia Care

**DOI:** 10.64898/2026.11.04.26362534

**Authors:** Brittyn Sutton, Isaiah Perry, Luke Kim, Anuhya Jujjuvarapu, Yoon Chae, Young-Tae Kim

**Author notes:** Correspondence and requests for materials should be addressed to Corresponding author: Young-Tae Kim, Ph.D. Department of Bioengineering 500 UTA Blvd ERB244 University of Texas at Arlington Arlington, TX 76010.

## Abstract

Falls and unmonitored patient movements remain leading causes of harm in healthcare settings. Current solutions – such as weight-based alarms, remote video monitoring, and in-person sitters – are reactive, labor-intensive, or prone to false alarms, often compromising both care quality and staff efficiency. We developed a machine learning-enabled, proactive-preventive patient monitoring platform that uses 17-keypoint pose estimation to detect changes in patient posture and generate alerts in real time. The Edge-AI device integrates a Raspberry Pi 5, Hailo-8L neural processing unit (NPU), and Raspberry Pi camera module 3, and runs Hailo-YOLOv8-Pose at 30 frames per second to classify postures (i.e., lying, Fowler’s, sitting, standing). Alerts are transmitted via Bluetooth to caregivers through earphones or speakers. System performance was evaluated across multiple scenarios, including bed and chair monitoring, free limb movement, partial body occlusion (e.g., blankets), and varying lighting and care environments. The platform maintained accurate monitoring under varying lighting conditions, including lights-off scenarios, and because all processing occurs locally without network dependence, it is designed to continue operating during network outages – an important resilience feature for care environments where connectivity may be interrupted. By abstracting images into skeletal keypoints without recording video, the system preserves privacy while potentially enabling earlier intervention and reducing false alarms and caregiver workload. This cost-effective, scalable system supports future enhancements such as dual alerting, multi-device full-room coverage, and continuous monitoring for dementia patients.

## Introduction

Patient safety is the highest priority in healthcare settings. Falls are a significant concern (e.g., 2 to 16 falls per 1,000 bed days with around 30% fall-related injuries) (Haines et al., 2008; Hester et al., 2016), often resulting from patients’ physical limitations or changes in mentation from their baseline due to conditions such as dementia (Allan et al., 2009; Fernando et al., 2017; Handelsman, 2011; Morris et al., 2022; van Dijk et al., 1993). To prevent falls, patient monitoring systems have been implemented, with bed alarm systems (e.g., Bed-Ex occupancy monitoring system) as the first line of defense. These systems, based on weight sensors, trigger an alarm when a change in weight is detected as the patient exits the bed (Lee et al., 2018; Shorr et al., 2012). Simultaneously, the bed’s interface communicates with third-party systems such as Vocera (Stryker) to sound external alarms, alerting nearby staff (Kuruzovich et al., 2008; Rumfield, 2021; Williams, 2008). However, current bed alarm systems have critical limitations. Because alarms are triggered only after a patient has exited the bed, the opportunity to intervene proactively is often missed – especially given patients’ compromised physical and/or mental conditions, where falls may already have occurred. Although beds allow sensitivity adjustments to address this issue, overly sensitive settings can cause frequent false alarms, leading to alarm fatigue – a major contributor to caregiver burnout (Asadi et al., 2022; Ding et al., 2023; Lewandowska et al., 2020; Solet & Barach, 2012; Storm & Chen, 2021). Additionally, alarms must be manually reset after activation; if not reset properly, monitoring lapses, further increasing the risk of falls. Other factors, such as improper bed calibration or the addition of equipment (e.g., oxygen tanks or other necessary medical equipment), can also compromise alarm effectiveness (Clodfelter, 2025; Winkler et al., 2025).

Other fall prevention methods include chair alarms (Knight et al., 2008), virtual sitters, and in-person patient sitters. Chair alarms operate similarly to bed alarms, sounding after the patient has already moved or fallen. Virtual sitters monitor patients remotely via cameras, alerting staff when necessary (Ergai et al., 2024; Quigley et al., 2021). However, remote monitoring cannot physically intervene to prevent harm and is limited by the number of available devices. In-person sitters, though effective, strain staffing resources and raise privacy concerns (Charles, 2023). Therefore, there is an urgent need for real-time, preventive patient monitoring technologies that improve fall prevention while preserving patient privacy and reducing caregiver workload.

In response to this need, we have developed a machine learning (ML)-enabled, real-time patient monitoring platform using a Hailo-8L neural processing unit (NPU) accelerator and Raspberry Pi. Our platform utilizes 17-keypoint pose estimation to continuously monitor patient posture in bed or chair environments at 30 frames per second (FPS), without storing or retaining identifiable video/images to protect privacy. By accurately detecting postures such as lying, Fowler’s position, sitting, and standing, the platform provides immediate voice alerts to caregivers via Bluetooth-paired earphones. To evaluate the platform’s patient monitoring and alert performance, four distinct patient monitoring trials were conducted: i) posture monitoring and alerting for lying, Fowler’s, sitting, and standing positions on a bed; ii) posture monitoring during free hand and leg motions on the bed; iii) posture detection when the patient’s body was partially covered by a blanket; and iv) monitoring sitting and standing transitions in a chair across various angles. We also assessed platform performance under different care environments, including varying locations and lighting conditions.

## Methods and Materials

### ML-enabled proactive-preventive patient monitoring platform

The proactive-preventive edge device consists of a Raspberry Pi 5 (64-bit, 8 GB RAM), a Hailo- 8L NPU (Hailo-8L AI chip: 13 trillion operations per second [TOPS] neural network inference accelerator), and a Raspberry Pi camera module 3 (Figure 1A). The assembled unit is mounted onto a 10-inch touchscreen monitor (Roadom), with the camera module attached using a 3D- printed, angle-adjustable mount (S_BELGI-3DELWORLD, 2022) (Figures 1B and 1C).

**Figure 1.**
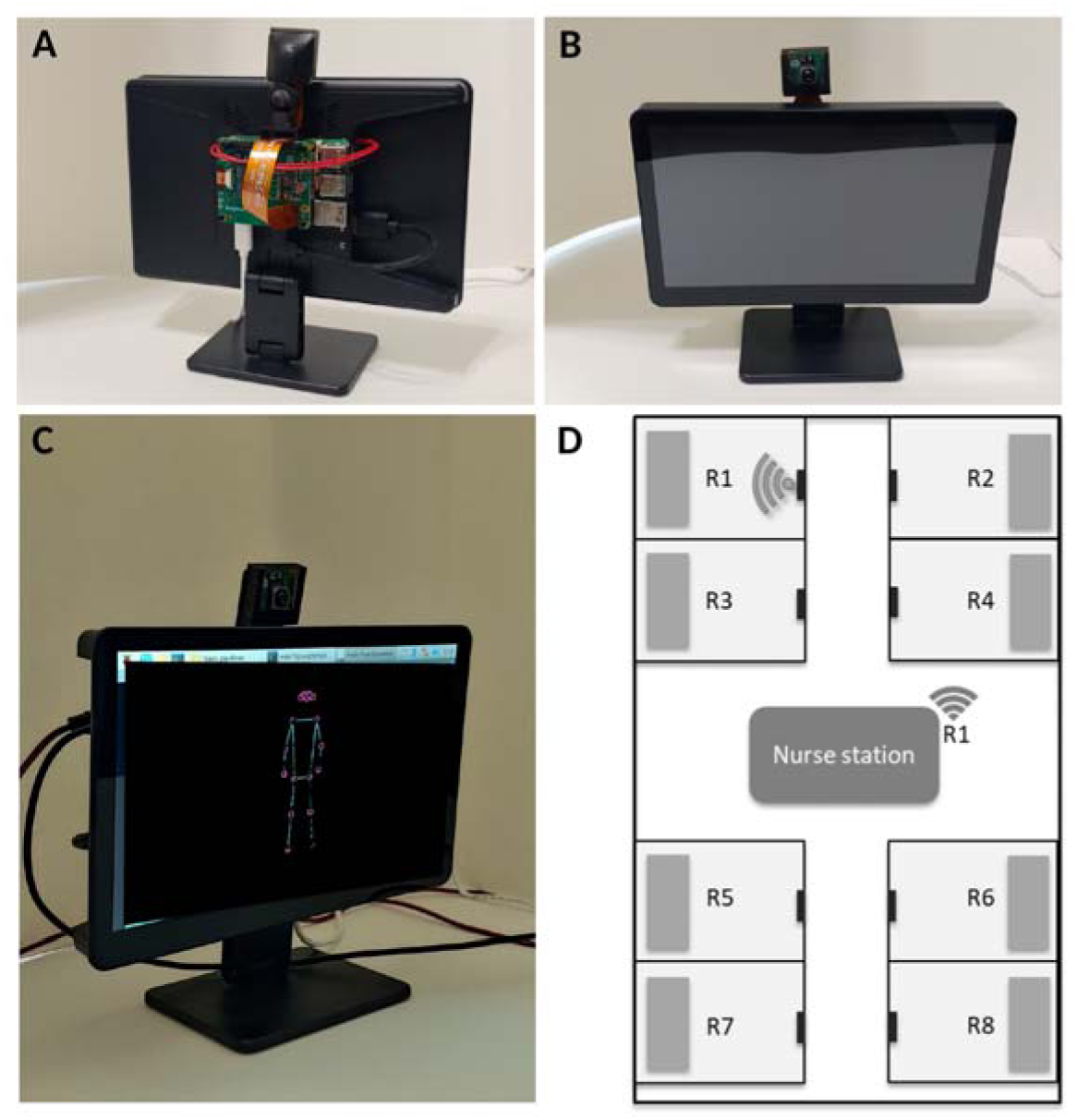
Edge-AI device for patient bed and chair monitoring. A-B) Real-time monitoring and alerting device mounted on a touchscreen display, with a 3D-printed, angle-adjustable camera holder. C) Touchscreen displaying the 17-keypoint skeleton of a subject standing on the floor; the on-screen camera image was removed to protect participant privacy, leaving only the skeletal overlay. D) Diagram of a clinical setting (e.g., nursing home: one or two caregivers and eight subjects lying in separate rooms; R1-R8). Black rectangles represent AI edge devices mounted on the wall of each subject’s room. The edge device in R1 detects a subject’s sitting motion and wirelessly informs the caregiver(s).

Figure 2 gives the hardware and communication block diagram. The camera is connected to the host over the CSI-2 interface; the Hailo-8L is attached over a single PCIe lane through the Raspberry Pi M.2 HAT. The host transfers the pre-processed frame tensor to the accelerator. Classification, alert-policy evaluation, and sound synthesis all run on the host CPU. The only signal that crosses the device boundary is the synthesized spoken label, streamed over Bluetooth to a caregiver earphone or bedside speaker.

**Figure 2.**
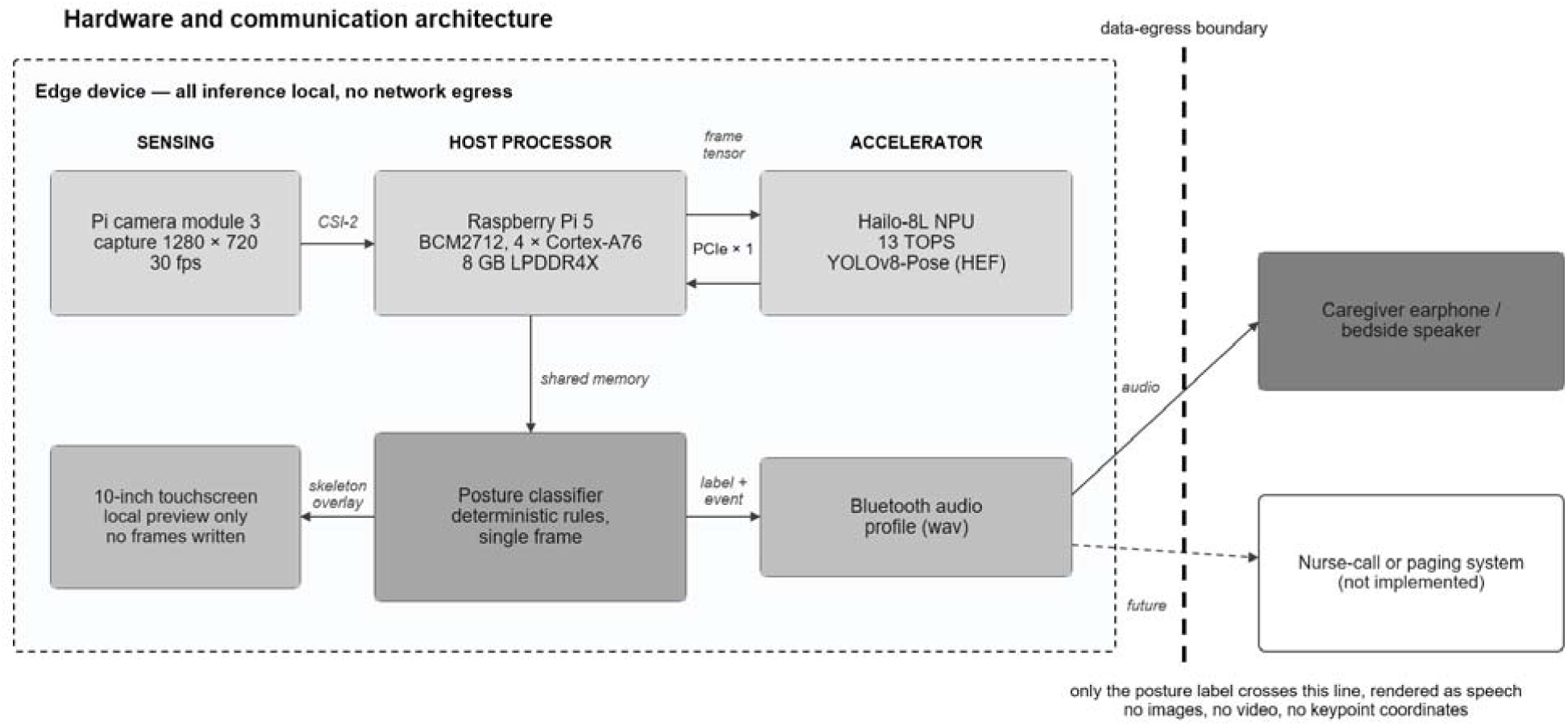
Edge-AI system architecture. Hardware and communication block diagram, showing the interfaces between camera, host processor, NPU accelerator, local display, and Bluetooth output; the data-egress boundary across which only a spoken posture label passes; and, as an unfilled box, the nurse-call or paging integration that is proposed but not implemented.

### 17-Keypoint human pose estimation for real-time monitoring and sound alerting

Pose estimation is performed using Hailo-YOLOv8-Pose, enabling 17-keypoint human pose detection at 30 FPS without excessive computational load. Lines are drawn connecting the detected keypoints, which include the nose, left and right eyes, ears, shoulders, elbows, wrists, hips, knees, and ankles (Figure 3). Upon detecting a person (with adjustable detection confidence thresholds), the X- and Y-coordinates of each keypoint are calculated and utilized to determine different postures (Figure 3E). Hardware setup and software installations followed established protocols (Hailo-AI, 2025; Raspberry Pi, 2024). Upon detecting programmed posture changes in bed or chair environments, the system delivers real-time voice alerts through Bluetooth-paired earphones or speakers.

**Figure 3.**
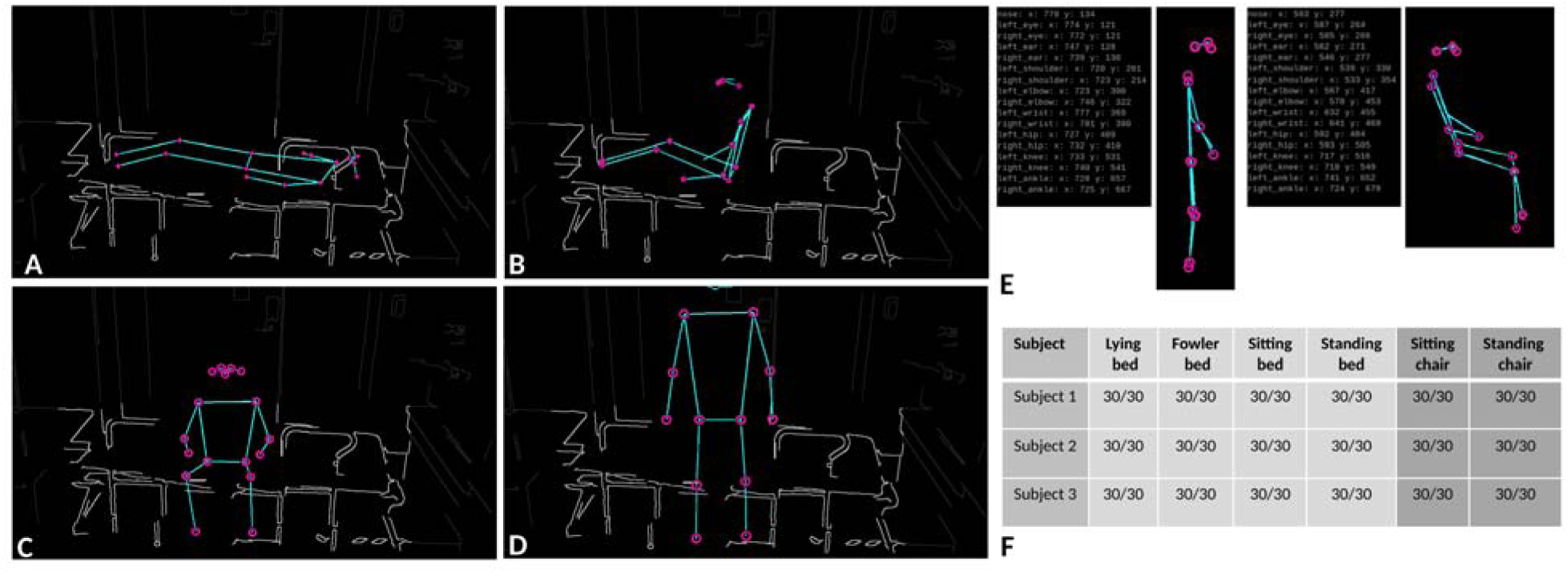
Monitoring of subject’s postures on a bed and chair. Representative 17-keypoint skeletons for four different postures: supine lying (A), Fowler’s position (B), sitting (C), and standing (D). Only the skeletal keypoint overlay generated by the device is shown; the camera image was removed to protect participant privacy. For context, the bed (light gray) and surrounding room (dark gray) are shown as line drawings traced from the original images; parts hidden behind the participant are omitted; panel E shows skeletons only. E) Screenshot of the X- and Y-coordinates of each keypoint corresponding to the subject in standing and sitting postures. F) Table summarizing three subjects’ posture detection and sound alerting on the bed and chair: 30/30 indicates 30 successful monitoring and sound alerts out of 30 trials.

### Monitoring of subject postures on bed

Four distinct postures on a bed – supine lying, Fowler’s position, sitting, and standing – were monitored and alerted (n=3 subjects; 30 trials per posture per subject; 90 trials per posture in total) (Figure 3). A custom Python program identified postures in real time based on the relative positions (X- and Y-coordinates; Figure 3E) of keypoints. When a subject remained in the same posture (e.g., lying on bed), a voice alert (“Subject is lying on bed”) was repeated every 10 seconds. The repeat interval can be adjusted based on situational needs. If the subject changed posture (e.g., from lying to Fowler’s), an immediate alert (“Subject is in Fowler’s position”) was issued to caregivers.

### Monitoring of subject postures on bed with free hand and leg motions

To assess robustness during natural movements, subjects’ postures were monitored and alerted while allowing free hand and leg motions (Figure 4). Natural movements included: supine lying with reversed body orientation; Fowler’s position with one or two hands resting on the mattress; sitting on the bed with one or two hands holding the bed rail; and sitting with one leg off the bed and one hand on the mattress.

**Figure 4.**
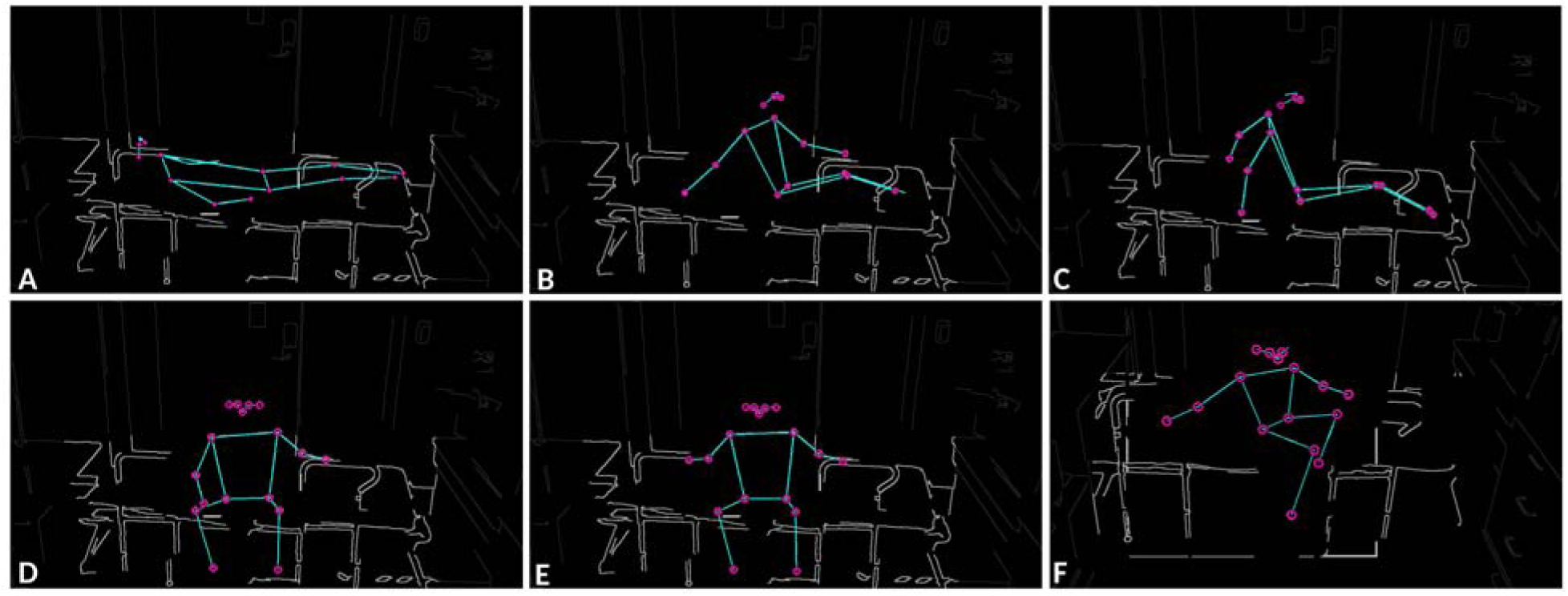
**Monitoring during free hand and leg motions on the bed**. Representative supine lying position with reversed body orientation (A). Fowler’s position with one (B) or both hands on mattress (C). Sitting on the bed with one (D) or both hands on the rail (E). Sitting on the bed with one leg out of the bed and one hand on the mattress (F). Only the skeletal keypoint overlay generated by the device is shown; the camera image was removed to protect participant privacy. For context, the bed (light gray) and surrounding room (dark gray) are shown as line drawings traced from the original images; parts hidden behind the participant are omitted.

### Monitoring of subject postures on bed covered by a blanket

To determine system performance when parts of the subject’s body were obscured, subjects’ postures were monitored and alerted while partially covered with a blanket (Figure 5). Tested scenarios included: lying with entire body covered except for the head; lying with shoulders exposed; lying with shoulders, hips, and both arms exposed; Fowler’s position with upper body exposed; and sitting with the upper body exposed while both hands rested on the knees or one hand on the rail.

**Figure 5.**
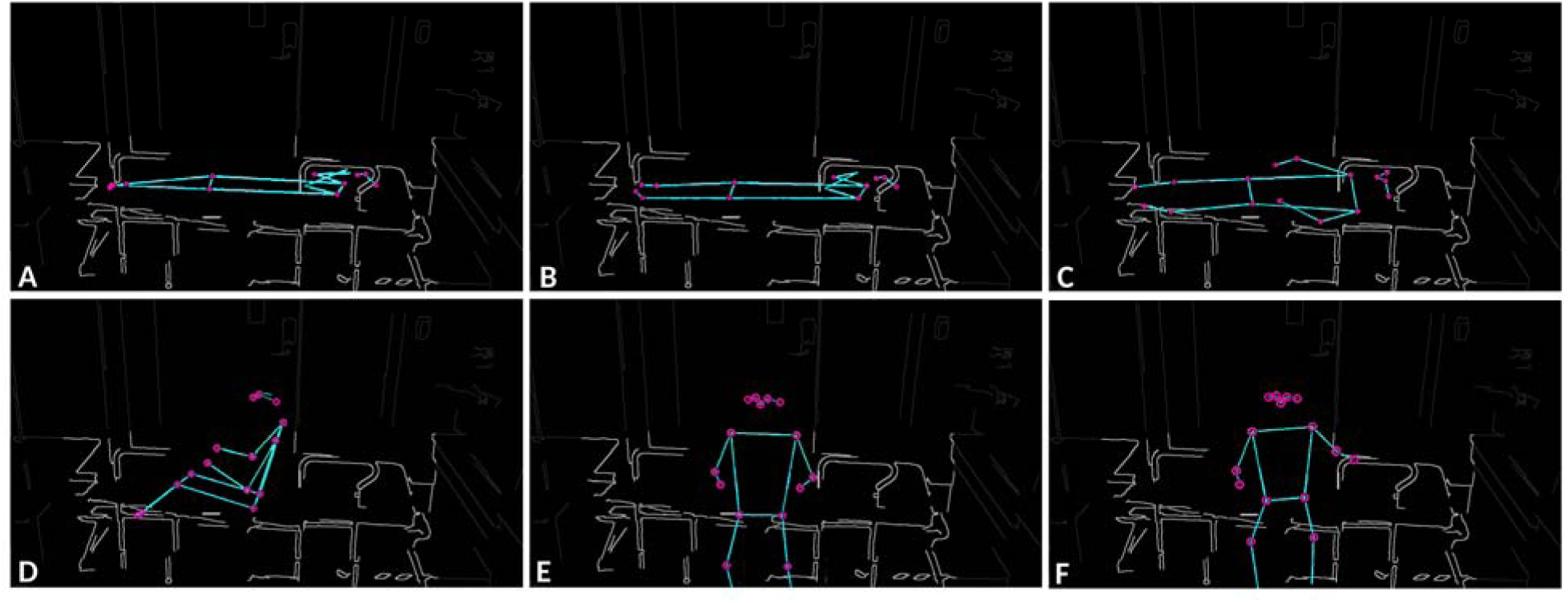
Monitoring of subject’s postures on the bed with a blanket. Representative lying with the entire body covered by a blanket except for the head (A). Lying with shoulders exposed (B). Lying with the shoulders, hips, and both arms exposed (C). Fowler’s position with the upper body exposed (D). Sitting with the upper body exposed and both hands on the knees (E) or one hand on the rail (F). Only the skeletal keypoint overlay generated by the device is shown; the camera image was removed to protect participant privacy. For context, the bed (light gray) and surrounding room (dark gray) are shown as line drawings traced from the original images; parts hidden behind the participant are omitted.

### Monitoring of subject postures on chair

Postures monitored on a chair included sitting (with various rotated positions) and standing (n=3 subjects; 30 trials per posture per subject; 90 trials per posture in total) (Figure 6). Similar to bed monitoring, if the subject maintained the same posture (e.g., sitting on chair), a voice alert (“Subject is sitting”) was repeated every 10 seconds. Upon posture change (e.g., sitting to standing), an immediate alert (“Subject is standing”) was issued to caregivers.

**Figure 6.**
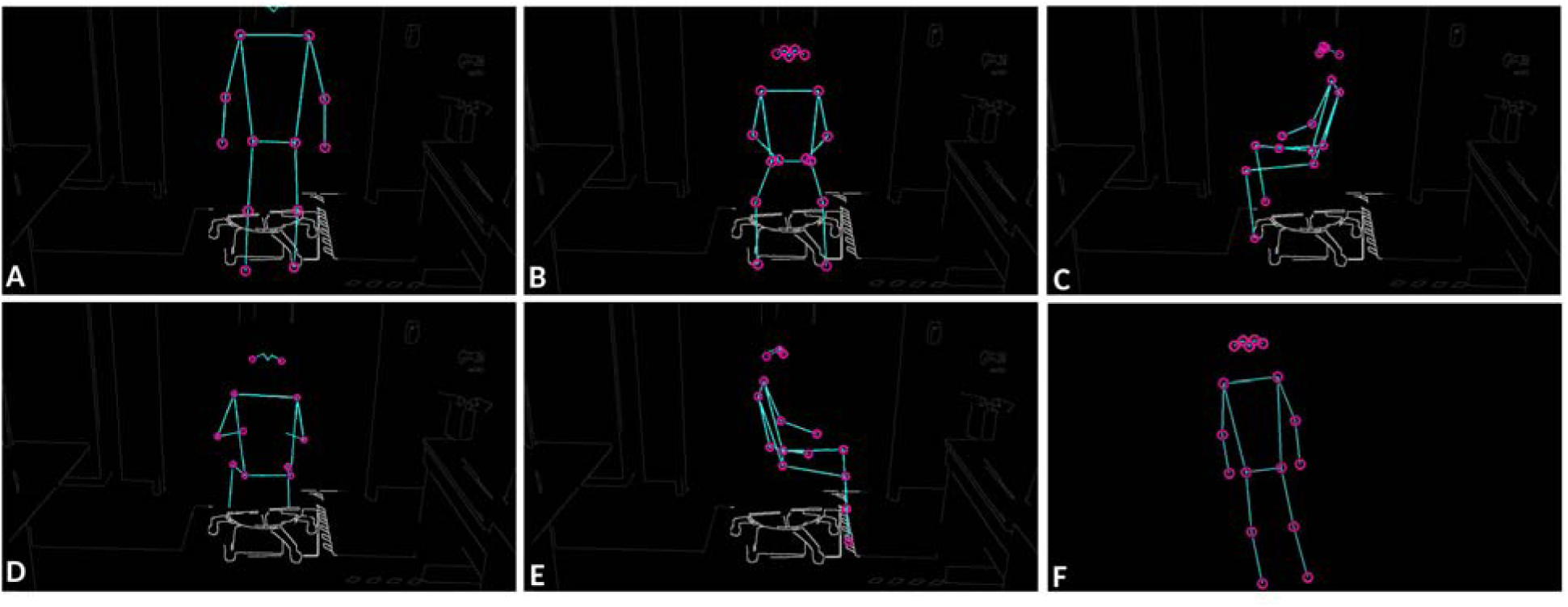
Monitoring of subject’s postures on a chair. Subject 1 in the laboratory environment: standing next to a chair (A), sitting facing the camera (B), sitting rotated 90 degrees (C), sitting rotated 180 degrees (D), and sitting rotated 270 degrees (E). Subject 2 standing in front of a chair in the home environment (F). Only the skeletal keypoint overlay generated by the device is shown; the camera image was removed to protect participant privacy. For context, in panels A–E the chair (light gray) and surrounding room (dark gray) are shown as line drawings traced from the original images; parts hidden behind the participant are omitted. Panel F (home environment) shows the skeleton only.

### Monitoring across different environmental conditions

To simulate realistic hospital and home care environments, monitoring performance was evaluated under: i) three different light brightness conditions: daytime, nighttime, and lights off and ii) two different testing locations: laboratory and home settings (Figure 7).

**Figure 7.**
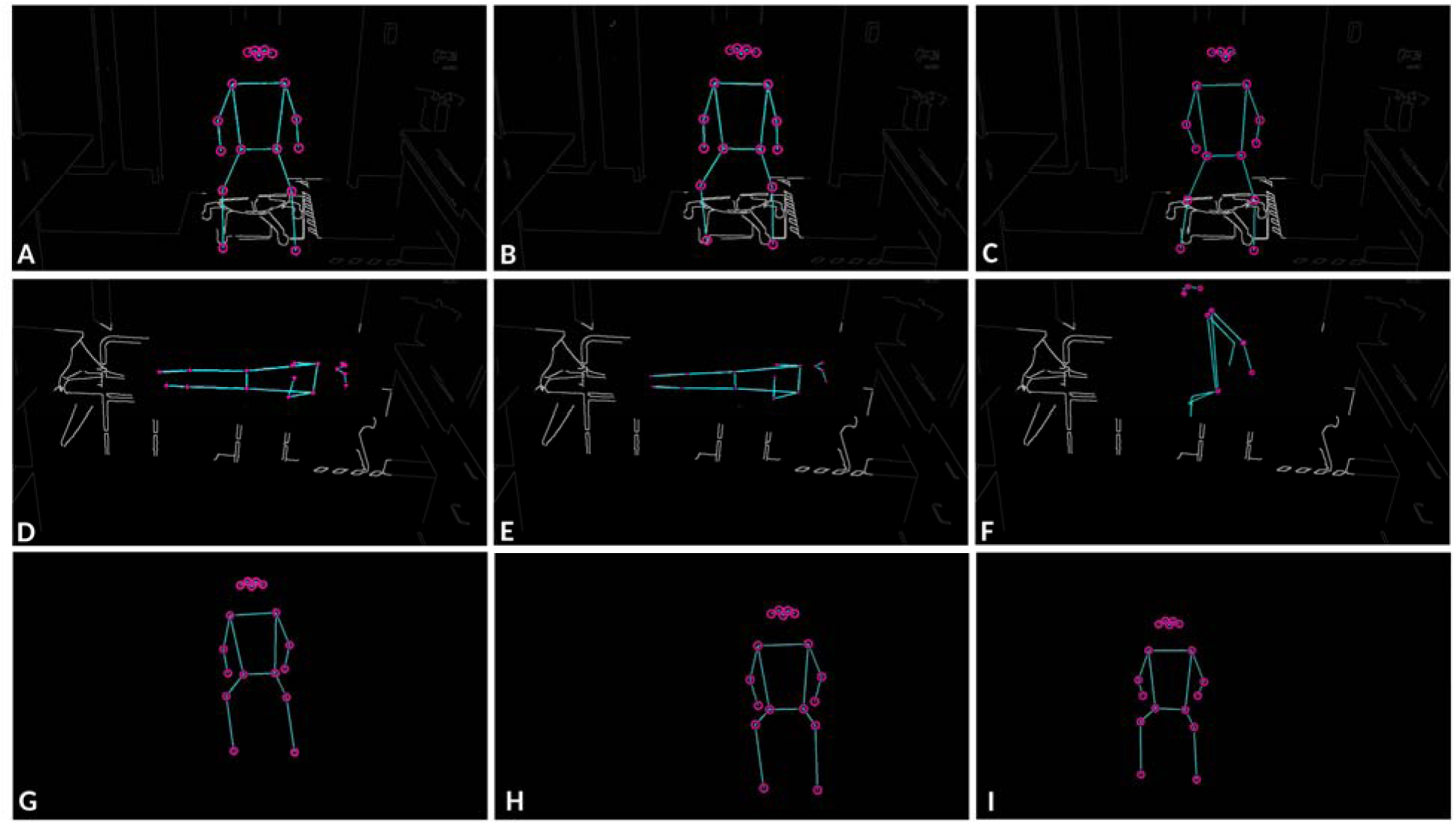
Monitoring of subject’s postures on a chair or bed under different environments: light brightness and location. Subject 1 sitting on a chair in the lab environment, with room light adjusted to daytime, nighttime, and lights off (A-C). Subject 1 on a bed covered with a blanket in the lab environment, with room light adjusted to daytime, nighttime, and lights off (D- F). Subject 2 sitting on a chair in the home environment, with room light adjusted to daytime, nighttime, and lights off (G-I). Only the skeletal keypoint overlay generated by the device is shown; the camera image was removed to protect participant privacy. For context, in panels A–F the chair or bed (light gray) and surrounding room (dark gray) are shown as line drawings traced from the original images, with parts hidden behind the participant omitted; for the lights-off panels (C, F), the outlines were traced from lit images of the same scene. Panels G–I (home environment) show the skeleton only.

## Results

### Real-time bed-posture monitoring and caregiver alerting

The ML-enabled patient monitoring platform detected and classified four different postures on the bed – lying, Fowler’s position, sitting, and standing – in 90/90 trials per posture (three subjects; 30 trials per posture per subject) (Figure 3). Four different postures were determined from the relative X- and Y-coordinates of 17 keypoints (Figure 3E). Real-time voice alerts were correctly triggered during posture transitions. When subjects maintained a single posture, repeated alerts were issued at the pre-programmed 10-second intervals without failure. Posture changes, such as from lying to Fowler’s or sitting to standing, were detected without perceptible delay, resulting in prompt voice notifications.

To ensure smooth performance, the real-time camera’s frame processing by the Hailo-8L NPU and audio playback by the Raspberry Pi 5 were handled asynchronously using separate threads.

This threading approach prevented audio playback from blocking real-time frame analysis. Additionally, a cooldown interval between successive audio outputs helped reduce redundant or excessive sound alerts.

### Monitoring during free hand and leg motions on bed

The edge platform demonstrated robust monitoring and alert performance when subjects performed free hand and leg movements while remaining in various postures (Figure 4). Postures such as reversed body orientation in lying position, one or two hands resting on the mattress during Fowler’s position, and partial bed-exiting postures were accurately monitored and classified. No misclassifications or missed alerts were observed in these scenarios, suggesting that the platform is robust to minor body movements.

### Monitoring of postures under blanket coverage

Monitoring performance was consistent even when subjects were partially covered with a blanket (Figure 5). In all tested covered postures – ranging from full body coverage (except for the head) to partial exposure of shoulders, hips, or arms – the platform reliably identified the correct posture and issued appropriate voice alerts. The blanket did not noticeably interfere with keypoint detection, indicating the platform’s robustness in common patient care environments where coverings are often used.

### Real-time monitoring and alerting of chair postures

Posture monitoring on a chair likewise detected and classified the postures in 90 of 90 trials per posture (three subjects; 30 trials per posture per subject), with appropriate sound alerts generated in all trials (Figure 3F). The platform accurately distinguished between sitting (even at various rotational angles) and standing postures across all three subjects (n=30 per subject) (Figure 6) and two different testing locations (lab and home environment). Consistent voice alerts were triggered every 10 seconds during prolonged sitting, and immediate alerts were issued upon standing transitions. No false alerts were observed during the trials.

### Monitoring under different lighting and location conditions

The platform’s posture detection and alerting performance remained stable under varying environmental conditions: i) various light conditions: Real-time posture recognition and alerts were consistent across daytime lighting, nighttime lighting, and even under lights-off conditions; and ii) various testing locations: Performance was equally robust in both laboratory and home environments, demonstrating the platform’s adaptability across different care settings (Figure 7). Across all tested conditions, the platform maintained real-time monitoring at 30 FPS, showed no noticeable degradation in posture detection accuracy, and reliably delivered Bluetooth voice alerts.

Importantly, because the Edge-AI device performs all processing locally and does not require external network connectivity, it is designed to continue posture monitoring during network outages; this was not tested experimentally in the present study. Such network independence could enhance patient safety by maintaining monitoring when hospital or home network connectivity is interrupted. Continuous operation during an electrical power outage, however, would additionally require a battery or uninterruptible power supply, which the current prototype does not include – a direction for future hardware development.

## Discussion

In this study, we developed an ML-enabled, proactive-preventive patient monitoring platform capable of real-time posture detection and alerting for subjects on beds or chairs in various care environments. The system leverages a lightweight Edge-AI device integrating a Raspberry Pi 5 and a Hailo-8L NPU to perform 17-keypoint human pose estimation. It delivers immediate posture alerts to caregivers through a Bluetooth-paired earphone or speaker, enabling proactive responses to patient movements.

The use of 17-keypoint pose estimation instead of full raw image data offers several key advantages in healthcare environments where privacy, real-time performance, and operational efficiency are critical: 1) Enhanced patient privacy: The system abstracts human figures into 17 skeletal keypoints, omitting facial details and identifiable features. This significantly reduces the risk of violating patient confidentiality. While subjects can view their posture status via the touchscreen interface (Figure 1C), no video recordings are stored, reducing privacy risks associated with stored video; 2) Real-time responsiveness: The Hailo-YOLOv8-Pose model runs at 30 FPS on the Hailo-8L NPU, enabling low-latency inference and instant alert delivery; 3) Lightweight data processing: The use of keypoint coordinates (X, Y positions of 17 keypoints; Figure 3E) reduces data bandwidth and processing needs compared to high-resolution video frames, optimizing system performance for real-time monitoring and alerts; 4) Lower power consumption: The pose-estimation-enabled monitoring operates efficiently within a 2–8 W range for the Hailo-8L NPU and 3–7 W for the Raspberry Pi 5 (Hailo-AI, 2024), making it suitable for always-on deployment in both clinical and homecare settings; 5) Scalability and modularity: Pose estimation outputs are easy to integrate with rule-based logic and alert systems, facilitating system expansion across multiple beds, rooms, or entire facilities without added complexity (Figure 1D); and 6) Suitable for monitoring patients in various care environments: Pose estimation is adjusted to only recognize persons so the monitoring is not disturbed by surrounding care environments such as home, hospital, bed, or chair types.

Compared to traditional monitoring approaches such as weight sensors, remote sitters, and in-person sitters, our ML-enabled platform offers several transformative advantages: 1) Proactive (proactive-preventive) monitoring: Unlike weight sensors, which typically trigger alarms only after a patient has exited the bed, our system detects and alerts on early posture transitions – such as sitting up or attempting to stand – giving caregivers an earlier window to respond than post-exit alarms allow. Because our evaluation did not include staged fall events, the platform’s effect on actual fall incidence remains to be established in future studies; 2) Reduced alarm fatigue: Traditional systems often produce false alarms due to minor patient movements such as rolling over or shifting weight. Our posture-aware alerts are context-specific, improving clinical response accuracy and reducing staff desensitization; 3) Privacy-conscious monitoring: Compared to remote sitters viewing live video or in-person sitters stationed bedside, the use of pose estimation preserves patient dignity and confidentiality through skeletal abstraction; 4) Operational efficiency and workforce support: ML-enabled monitoring alleviates the staffing burden, offering consistent and uninterrupted surveillance across shifts. The system is unaffected by fatigue, distractions, or turnover – challenges inherent to human sitters; and 5) Robust performance across settings: The system is resilient to variability in lighting, room configurations, and furniture types, areas where traditional solutions often require recalibration or fail altogether.

To further advance the capabilities of this first-generation ML-based platform, we propose several future enhancements: First, dual sound alerting: In addition to caregiver alerts delivered via Bluetooth, the system could directly communicate with patients. Upon detecting posture transitions (e.g., from lying to Fowler’s position or sitting to standing), the device could issue immediate verbal prompts – such as “Please stay in bed” or “Help is on the way” – through built-in or paired speakers, enhancing patient safety and compliance; second, full-room monitoring using multiple devices: As camera-based systems are limited to their field of view, deploying multiple synchronized units can eliminate blind spots. With a unit fabrication cost below $200, a multi-device setup is feasible for full-room coverage without compromising affordability; and third, automatic detection of multi-person presence: To maintain data integrity and privacy, the system can be programmed to temporarily pause monitoring when additional individuals (e.g., caregivers) are detected in the frame. It can then notify out-of-room staff (e.g., “Monitoring temporarily paused due to caregiver presence”), and resume automatically once the room is cleared (e.g., “Monitoring resumed for Room 1”) (Supplementary Figure 1).

**Supplementary Figure 1.**
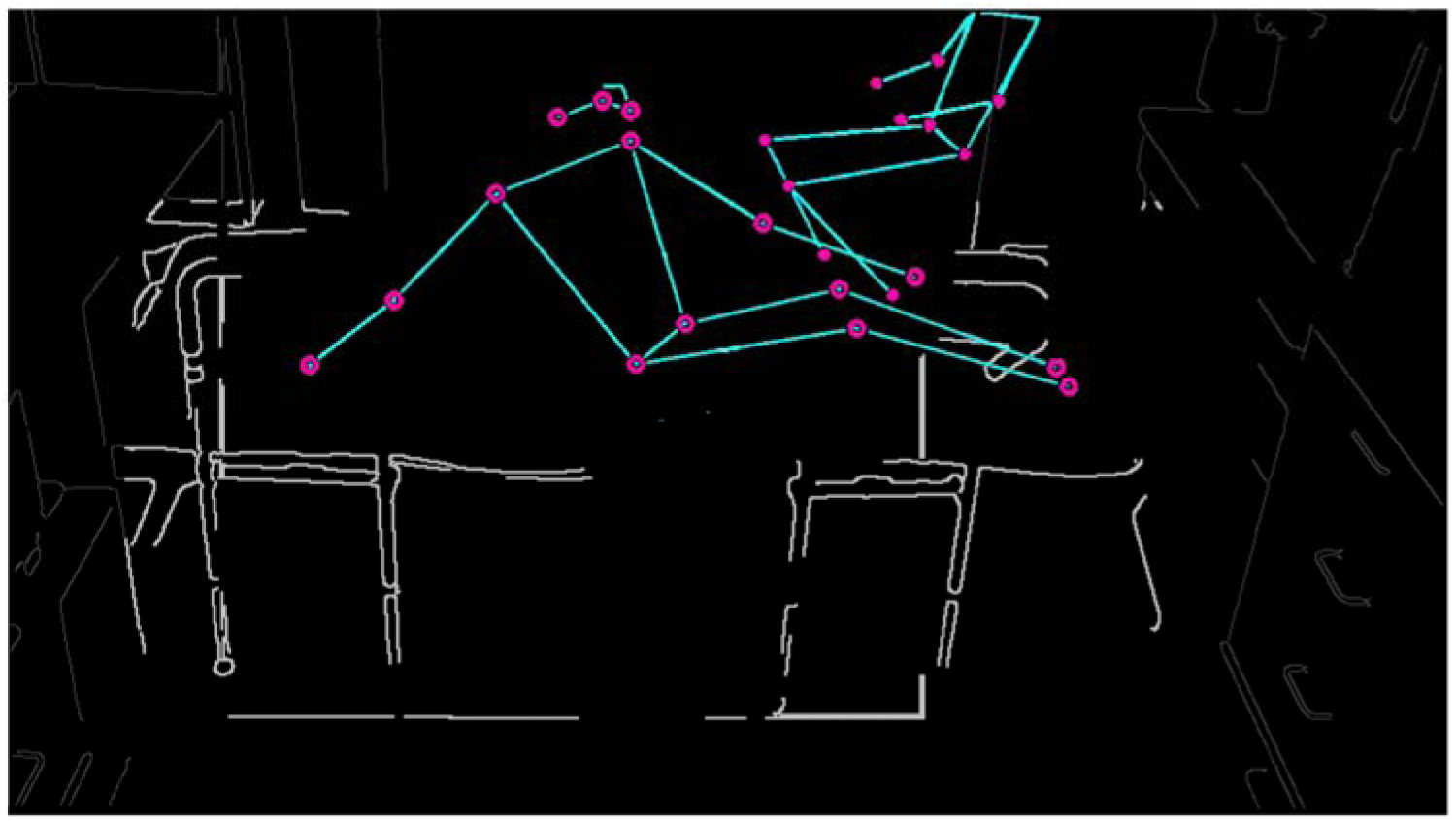
Subject lying on a bed with a caregiver. Representative image showing two persons in the camera view: a subject lying on a bed and a caregiver sitting on a chair. Only the skeletal keypoint overlay generated by the device is shown; the camera image was removed to protect participant privacy. For context, the bed (light gray) and surrounding room (dark gray) are shown as line drawings traced from the original images; parts hidden behind the participants are omitted.

Finally, Supplementary Figure 2 highlights that these Edge-AI devices can be assembled into highly compact, screenless form factors. This compact footprint enables discreet placement on patient beds, wheelchairs, and bedside furniture, allowing continuous real-time monitoring without interfering with clinical workflow or patient comfort. Together, these enhancements illustrate the potential of this platform to evolve into a robust, patient-centered, and privacy- preserving monitoring ecosystem.

This study has several limitations. The evaluation involved a small sample of three subjects, all of whom were also co-authors, rather than an independent cohort of older adults or individuals with dementia; performance across different body types, mobility patterns, and comorbidities remains to be established. The study was reviewed by the Office of Regulatory Services, Division of Research Administration, The University of Texas at Arlington, which determined that the activity constituted Not Human Subjects Research (Non-HSR) and that Institutional Review Board review and approval were not required. Future studies involving external participants, particularly vulnerable populations such as individuals with dementia, will require prospective ethics/IRB review and appropriate consent procedures. The evaluation assessed posture and posture-transition classification but did not include staged falls or near-fall events, so whether earlier alerts translate into fewer actual falls or faster caregiver response has not been tested. The posture classifier relies on fixed geometric thresholds rather than a model trained on various postures and body types, which may limit generalizability beyond the tested conditions. Finally, all trials were conducted in a controlled laboratory or single home setting; performance in busier clinical environments with multiple occupants, medical equipment, or complex bedding was not assessed. Larger, IRB-approved studies with independent participants and real-world deployment are needed to validate the platform’s fall-prevention benefit.

**Supplementary Figure 2.**
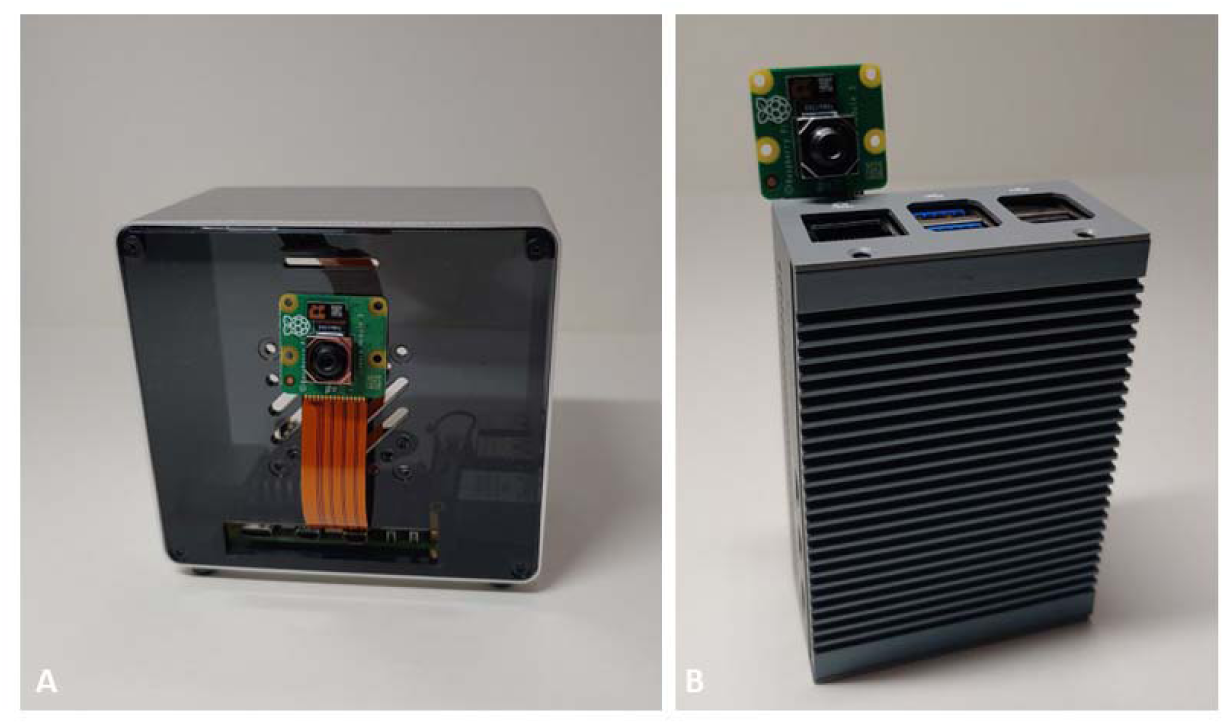
Compact, screenless preventive Edge-AI devices for patient bed and chair monitoring. A Raspberry Pi 5, a Hailo-8L NPU, and a Raspberry Pi camera module 3 can be assembled into compact form factors without a screen: 10 x 9 x 6 cm (A) and 9 x 6 x 3 cm (B).

## Conclusion

This study presents the development and feasibility evaluation of a machine learning-enabled, proactive-preventive patient monitoring and alerting platform using 17-keypoint human pose estimation for real-time posture detection on beds and chairs across various care environments. Implemented on a low-cost Edge-AI device integrating a Raspberry Pi 5 and a Hailo-8L NPU, the system delivers immediate, privacy-conscious alerts to caregivers via Bluetooth-paired speakers or earphones. Unlike traditional systems such as weight sensors or in-person sitters, this platform enables proactive detection of posture transitions – including early indicators of bed exit, such as sitting up or moving to the bed edge – and operates reliably under the lighting, room, and coverage conditions tested here.

The results demonstrate high performance and robustness in detecting a wide range of postures, including those partially obscured by blankets or involving free limb movements.

These findings support the system’s potential as a scalable, always-on, and privacy-preserving solution for continuous patient monitoring. Future enhancements such as dual sound alerts, multi-device room coverage, and automated interruption handling will further strengthen its utility in clinical and homecare settings. Overall, the proposed platform represents a significant step toward intelligent, preventive, and cost-effective patient safety monitoring, and it can be further adapted to support dementia patient care through continuous mobility and behavioral monitoring.

## Statements and Declarations Consent to Participate

All individuals who performed the predefined postures were investigators/authors of the study and voluntarily agreed to participate in the technical feasibility testing.

## Ethics Statement

This study was reviewed by the Office of Regulatory Services, Division of Research Administration, The University of Texas at Arlington, Arlington, Texas, USA. The reviewing body determined that this study constitutes Not Human Subjects Research (Non-HSR) and that Institutional Review Board review and approval were not required. The study was limited to technical feasibility testing of the Edge-AI posture-monitoring device. All individuals who performed the predefined postures were investigators/authors of the study. Continuous RGB video was processed in real time and was not recorded or retained as study data. Representative still images were captured solely to illustrate device operation. In accordance with medRxiv policy, the figures in this preprint show only the device-generated skeletal keypoint overlays, with all camera image content removed (for context, the bed, chair, and laboratory room are shown as line drawings traced from the original images, excluding the participants); the original images are available from the corresponding author upon reasonable request. For performance analysis, only AI-generated skeletal/pose-estimation outputs, posture classifications, and device detection/alert outcomes were retained. No clinical, physiological, behavioral, usability, or other person-level outcomes were collected or analyzed.

## **Clinical trial number**: not applicable

## Availability of data and materials

The datasets used and/or analyzed during the current study, including representative images of device operation, are available from the corresponding author on reasonable request.

## Author Contributions

Y.T.K. was the principal investigator. B.S., Y.C., and Y.T.K. wrote the paper. Y.T.K. and B.S. developed the idea and designed the experiments. L.K. coded and fabricated the Edge-AI device. A.J. adapted and 3D-printed the camera holder. Y.T.K., B.S., I.P., L.K., and A.J. carried out all experiments and produced all data.

## Data Availability

All data produced in the present study are available upon reasonable request to the authors

## Acknowledgements

Not applicable.

## References

Allan, L. M., Ballard, C. G., Rowan, E. N., & Kenny, R. A. (2009). Incidence and prediction of falls in dementia: A prospective study in older people. PLoS One, 4(5), e5521. 10.1371/journal.pone.0005521

Asadi, N., Salmani, F., Asgari, N., & Salmani, M. (2022). Alarm fatigue and moral distress in ICU nurses in COVID-19 pandemic. BMC nursing, 21(1), 125.

Charles, M. (2023). Implementing a Virtual Sitter Program to Reduce Patient Falls and Cost. Doctor of Nursing Practice project, The University of Alabama in Huntsville.

Clodfelter, A. D. (2025). Improving Alarm Management Practices: Wireless Bed Exit Alerts on Medical-Surgical Units. CIN: Computers, Informatics, Nursing, 43(8), e01324. 10.1097/CIN.0000000000001324

Ding, S., Huang, X., Sun, R., Yang, L., Yang, X., Li, X.,…Wang, X. (2023). The relationship between alarm fatigue and burnout among critical care nurses: A cross sectional study. Nursing in critical care, 28(6), 940–947.

Ergai, A., Spiva, L., Thurman, S., Hatfield, M., McCollum, M., & Holmes, M. (2024). The effectiveness of remote video monitoring on fall prevention and nurses’ acceptance. Journal of Nursing Care Quality, 39(1), 24–30.

Fernando, E., Fraser, M., Hendriksen, J., Kim, C. H., & Muir-Hunter, S. W. (2017). Risk Factors Associated with Falls in Older Adults with Dementia: A Systematic Review. Physiother Canada, 69(2), 161–170. 10.3138/ptc.2016-14

Hailo-AI. (2024). Hailo-8 Power consumption. https://community.hailo.ai/t/hailo-8-power-consumption/2879

Hailo-AI. (2025). Hailo-rpi5-examples. https://github.com/hailo-ai/hailo-rpi5-examples

Haines, T., Kuys, S. S., Morrison, G., Clarke, J., & Bew, P. (2008). Balance impairment not predictive of falls in geriatric rehabilitation wards. The Journals of Gerontology Series A: Biological Sciences and Medical Sciences, 63(5), 523–528.

Handelsman, J. (2011). Falls among older adults: A public health concern. Perspectives on Hearing Conservation and Occupational Audiology, 12(1), 13–18.

Hester, A. L., Tsai, P.-F., Rettiganti, M., & Mitchell, A. (2016). CE: original research: predicting injurious falls in the hospital setting: implications for practice. AJN The American Journal of Nursing, 116(9), 24–31.

Knight, H., Lee, J.-K., & Ma, H. (2008). Chair alarm for patient fall prevention based on gesture recognition and interactivity. *In* 2008 30th Annual International Conference of the IEEE Engineering in Medicine and Biology Society (pp. 3698–3701). IEEE.

Kuruzovich, J., Angst, C. M., Faraj, S., & Agarwal, R. (2008). Wireless communication role in patient response time: a study of Vocera integration with a nurse call system. *CIN: Computers, Informatics*, Nursing, 26(3), 159–166.

Lee, C.-N., Yang, S.-C., Li, C.-K., Liu, M.-Z., & Kuo, P.-C. (2018). Alarm system for bed exit and prolonged bed rest. *In* 2018 International Conference on Machine Learning and Cybernetics (ICMLC) (pp. 439–443). IEEE.

Lewandowska, K., Weisbrot, M., Cieloszyk, A., Mędrzycka-Dąbrowska, W., Krupa, S., & Ozga, D. (2020). Impact of alarm fatigue on the work of nurses in an intensive care environment—a systematic review. International journal of environmental research and public health, 17(22), 8409.

Morris, M. E., Webster, K., Jones, C., Hill, A.-M., Haines, T., McPhail, S., Kiegaldie, D., Slade, S., Jazayeri, D., Heng, H., Shorr, R., Carey, L., Barker, A., & Cameron, I. (2022). Interventions to reduce falls in hospitals: a systematic review and meta-analysis. Age and Ageing, 51(5), afac077. 10.1093/ageing/afac077

Quigley, B. H., Renz, S. M., & Bradway, C. (2021). Fall prevention and injury reduction utilizing virtual sitters in hospitalized patients: a literature review. CIN: Computers, Informatics, Nursing, 39(12), 929–934.

Raspberry Pi. (2024). *How to set up the Raspberry Pi AI Kit with Raspberry Pi 5*. Raspberry Pi. https://www.raspberrypi.com/news/how-to-set-up-the-raspberry-pi-ai-kit-with-raspberry-pi-5

Rumfield, J. D. (2021). The Association Between Patient Falls and Wired and Wireless Patient Communication Systems [Doctoral dissertation, Trident University International].

S_BELGI-3DELWORLD. (2022). Raspberry Pi camera holder (DUCKLING). cults3d.com. https://cults3d.com/en/3d-model/tool/raspberry-pi-camera-holder-duckling-camera-holder-for-octoprint

Shorr, R. I., Chandler, A. M., Mion, L. C., Waters, T. M., Liu, M., Daniels, M. J.,…Miller, S. T. (2012). Effects of an intervention to increase bed alarm use to prevent falls in hospitalized patients: a cluster randomized trial. Annals of internal medicine, 157(10), 692–699.

Solet, J. M., & Barach, P. R. (2012). Managing alarm fatigue in cardiac care. Progress in Pediatric Cardiology, 33(1), 85–90.

Storm, J., & Chen, H. C. (2021). The relationships among alarm fatigue, compassion fatigue, burnout and compassion satisfaction in critical care and step down nurses. Journal of clinical nursing, 30(3-4), 443–453.

van Dijk, P. T., Meulenberg, O. G., van de Sande, H. J., & Habbema, J. D. (1993). Falls in dementia patients. The Gerontologist, 33(2), 200–204. 10.1093/geront/33.2.200

Williams, J. S. (2008). Vocera voice badges cut response times. Biomedical Instrumentation & Technology, 42(3), 193–194.

Winkler, A., Pallauf, M., Krutter, S., Kutschar, P., Osterbrink, J., & Nestler, N. (2025). Sensor- based prevention of falls and pressure ulcers: A scoping review. International Journal of Medical Informatics, 199, 105878.

